# Safety and effectiveness of SGLT2-inhibitors in people with type 2 diabetes over 70: UK population-based study using an Instrumental Variable approach

**DOI:** 10.1101/2024.01.04.24300832

**Authors:** Laura Maria Güdemann, Katie G. Young, Nicholas J. M. Thomas, Rhian Hopkins, Robert Challen, Angus G. Jones, Andrew T. Hattersley, Ewan R Pearson, Beverley M. Shields, Jack Bowden, John M. Dennis, Andrew P. McGovern, the Mastermind consortium

## Abstract

**Objective:** Older adults are underrepresented in trials, meaning the benefits and risks of glucose lowering agents in this age group are unclear. We applied causal analysis to assess the safety and effectiveness of SGLT2-inhibitors in people with type 2 diabetes (T2D) over 70.

**Research Design and Methods:** Hospital-linked UK primary care data (Clinical Practice Research Datalink, 2013-2020) were used to compare adverse events and effectiveness in individuals initiating SGLT2-inhibitors compared to DPP4-inhibitors. Analysis was age-stratified: <70 years (SGLT2-inhibitors n=66810, DPP4-inhibitors n=76172), ≥70 years (SGLT2-inhibitors n=10419, DPP4-inhibitors n=33434). Outcomes were assessed using the Instrumental Variable causal inference method and prescriber preference as instrument.

**Results:** Risk of DKA was increased with SGLT2-inhibitors in those aged ≥70 (Incidence risk ratio compared to DPP4i: 3.82 [95%CI 1.12,13.03]), but not in those <70 (1.12 [95%CI 0.41,3.04]). However incidence rates with SGLT2-inhibitors in those ≥70 was low (29.6 [95%CI 29.5,29.7]) per 10000 person-years. SGLT2-inhibitors were associated with similarly increased risk of genital infection in both age groups (IRR <70 2.27 [2.03,2.53]; ≥70 2.16 [1.77,2.63]). There was no evidence of an increased risk of volume depletion, poor micturition control, urinary frequency, falls or amputation with SGLT2-inhibitors in either age group. In those ≥70, HbA1c reduction was similar with SGLT2-inhibitors and DPP4-inhibitors (−0.3 mmol/mol [−1.6,1.1], −0.02% [0.1,0.1]), but in those <70 SGLT2-inhibitors were more effective (−4 mmol/mol [4.8,−3.1], −0.4% [−0.4,−0.3]).

**Conclusions:** Causal analysis suggests SGLT2-inhibitors are effective in adults ≥70, but increase risk for genital infections and DKA. Our study extends RCT evidence to older adults with T2D.

**Article Highlights:** Why did we undertake this study?

– Current guidelines for type 2 diabetes recommend an individualised approach to treatment, but evidence for older adults is limited.

What is the specific question(s) we wanted to answer?

– To assess the safety and effectiveness of SGLT2-inhibitors in older adults by applying a causal inference framework to address potential confounding bias in observational data.

What did we find?

– SGLT2-inhibitors are effective in reducing HbA1c and weight and generally safe for older adults. Adverse events in this older group include genital infections and a small increase in DKA.

What are the implications of our findings?

– SGLT2-inhibitors are effective and safe for older adults, but clinicians should be aware of the risks for genital infections and DKA.

## Introduction

Current type 2 diabetes (T2D) guidelines recommend an individualised approach to treatment that takes into account preferences, comorbidities, risks from polypharmacy, and the likelihood of long-term benefit from interventions, [1, 2] but clear guidance on therapeutic strategies for the management of T2D in older adults is limited. [3] For older adults, specific treatment considerations are likely to be needed, due to increased comorbidities, age-related changes in physiology and pharmacodynamics, as well as possible increased propensity to adverse medication effects.

Under current guidelines, a large proportion of older people with T2D would be recommended SGLT2-inhibitors due to their cardiorenal benefits, and irrespective of their glycaemic control [1, 4]. SGLT2-inhibitors have well described benefits, particularly cardiorenal and the promotion of weight loss [5, 6, 7, 8], but also possible risks, which may limit their use for older people. [3] Well-established risks of SGLT2-inhibitors are genital infections and due to their mode of action, volume depletion is possible. [6, 9] These side effects could be of particular concern for older adults where incontinence, dehydration and dizziness could have more severe consequences compared with a younger population. [10, 11, 12, 13] Additionally, dehydration or dizziness can also lead to falls in older people. [14] Further adverse events of concerns of SGLT2-inhibitors are lower limb amputations [9]. Reports of possible association of SGLT2-inhibitors and diabetic ketoacidosis (DKA) has prompted the FDA [15] and the EMA [16] to issue warnings. Older people may also present with more frequent acute complications, such as infections, which are additional risk factors of DKA. [17]

In order to develop targeted guidelines for the management of T2D in older adults, evidence on risks and benefits of treatments in this age group is needed. [3] However, older people are underrepresented in randomized clinical trials (RCTs) and caution is needed when extrapolating RCT evidence for this group. [3, 18] Observational studies of the older T2D population have the potential to provide insights that are not provided by RCTs. Previous post-hoc RCT analyses [13, 19, 20, 21] have examined risks in older adults, but have very small sample sizes for older people with T2D, and therefore might suffer from outlier effects. [13] Also, without detailed data on characteristics, comorbidities and concomitant medications the results from observational studies may be affected by unmeasured confounding which can bias treatment effect results. [14]

We therefore aimed to examine the relative risks and benefits of SGLT2-inhibitors in older people compared to DPP4-inhibitors using large-scale routine primary and linked secondary care data. We employ an Instrumental Variable approach, exploiting systematic variation in practitioners’ prescribing preference as the instrument, to estimate the impact of receiving SGLT2-inhibitors compared to DPP4-inhibitors on a range of adverse events and important treatment outcomes, analogous to a randomised controlled trial.

## Methods

### Study design and participants

In this retrospective cohort study, UK routine primary care data were accessed from Clinical Research Datalink (CPRD) Aurum (October 2020 download). CPRD is a UK representative sample covering approximately 13% of the population in England. [22] CPRD Aurum was linked to Hospital Episode Statistics (HES), Office for National Statistics (ONS) death registrations and individual-level Index of Multiple Deprivation (IMD). Individuals with T2D were identified according to a previously published protocol [23] based on the presence of a diagnostic code for diabetes and the prescription of one or more glucose lowering medications. Type 1 diabetes and other types of diabetes were excluded. [23] The analysis included new users of SGLT2-inhibitors (Canagliflozin, Dapagliflozin, Empagliflozin, Ertugliflozin), initiating treatment after 1^st^ January 2013 and with an identifiable date of T2D diagnosis. The comparison cohort was new users of DPP4-inhibitors (Alogliptin, Linagliptin, Sitagliptin, Saxagliptin, Vildagliptin), as these agents represent the most commonly prescribed drug class after metformin in the UK, and have no known association with the SGLT2-inhibitors-associated adverse events of interest evaluated in this study. All available follow-up data was considered in the analysis up to the point of data extraction. Individuals with a baseline HbA1c outside of the range 53-120 mmol/mol (7% - 13.1%) were excluded from the analysis, reflecting on the threshold for glucose-lowering medication initiation in clinical guidelines and severe hyperglycemia. Additionally, individuals with renal impairment indicated with a glomerular filtration rate (eGFR) of less than 45 mL/min/1.73 m^2^ were excluded, as SGLT2-inhibitors were not licensed for use below this threshold for the majority of the study period. Further exclusion criteria are summarized in Figure 1. Our cohort was split into a population aged less than 70 years at treatment initiation and an older population (≥70 years).

**Figure 1.**
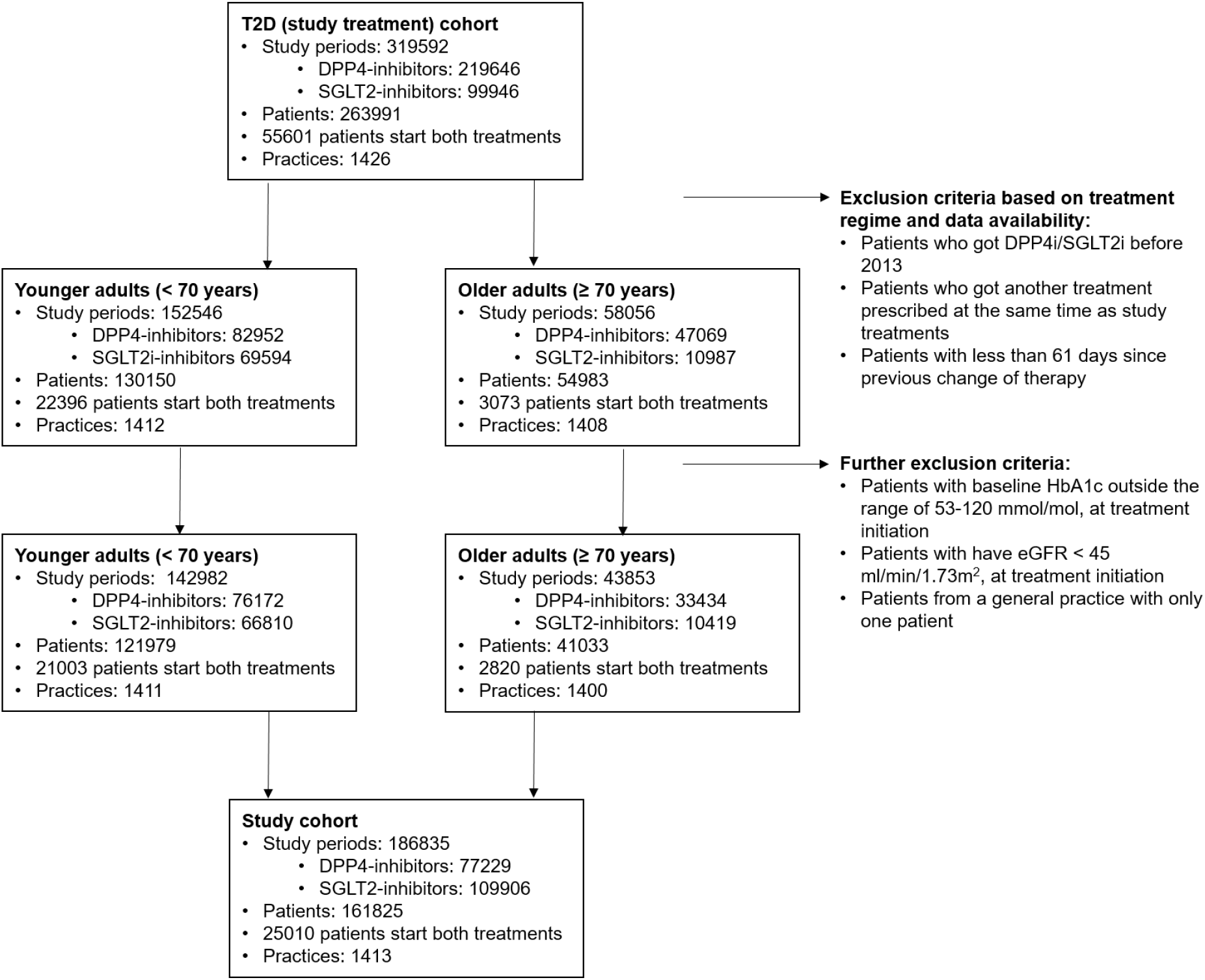
Flow chart of study cohort selection, age-stratified.

### Outcomes

Adverse events (AE) included in the analysis were genital infections, micturition control (urge incontinence, urgency, stress incontinence, or nocturnal enuresis), volume depletion and dehydration, urinary frequency, falls, lower limb amputation and DKA. The occurrence of each AE was measured up to 3 years after treatment initiation and censoring of the follow-up time was implemented in case of a discontinuation of the study treatment or start of the comparison study treatment. Individuals were therefore followed up until the earliest of: date of the outcome of interest, discontinuation of the study treatment, start of comparison study treatment, date of practice deregistration/death, end of study period, or 3 years. Occurrences of AEs were identified using diagnosis code lists published at: https://github.com/Exeter-Diabetes/CPRD-Codelists. Genital infections were identified with either a diagnosis code for a specific genital infection (e.g. candida vaginitis or vulvo-vaginitis in women, balanitis, balanoposthitis in men), a prescription for antifungal therapy used specifically to treat genital infections (e.g. an antifungal vaginal pessary), or a non-specific diagnosis of “thrush” with a topical antifungal prescribed on the same day. [24] The diagnosis codes to identify amputation AEs were taken from Pearson-Stuttard et al. [25]. DKA was identified using HES hospitalization data. Treatment outcomes to assess relative effectiveness of SGLT2-inhibitors included achieved glycated haemoglobin (HbA1c in mmol/mol and %) and weight (kg) on unchanged therapy. These outcome measurements were taken as the closest recorded value to 12 months post treatment initiation, within a window of 3 to 15 months.

### Covariates

Measured covariates for all outcome models were extracted following our previous protocol [23] together with general information about individuals, including sociodemographic features (age, sex, ethnicity and deprivation) and treatment history, important biomarkers as well as history of relevant comorbidities. Biomarker baseline values are defined nearest to treatment initiation up to 2 years before and 7 days after initiation. Initiation of relevant additional treatments such as diuretics, have been observed up to 3 months before treatment initiation and comorbidities have been characterised to be within 1 year, 1-5 years or >5 to treatment initiation. A summary of all covariates is given in Table 1; a cohort description and a comprehensive overview of the biomarker and comorbidity definitions are given here: https://github.com/Exeter-Diabetes/CPRD-Cohort-scripts.

**Table 1:**
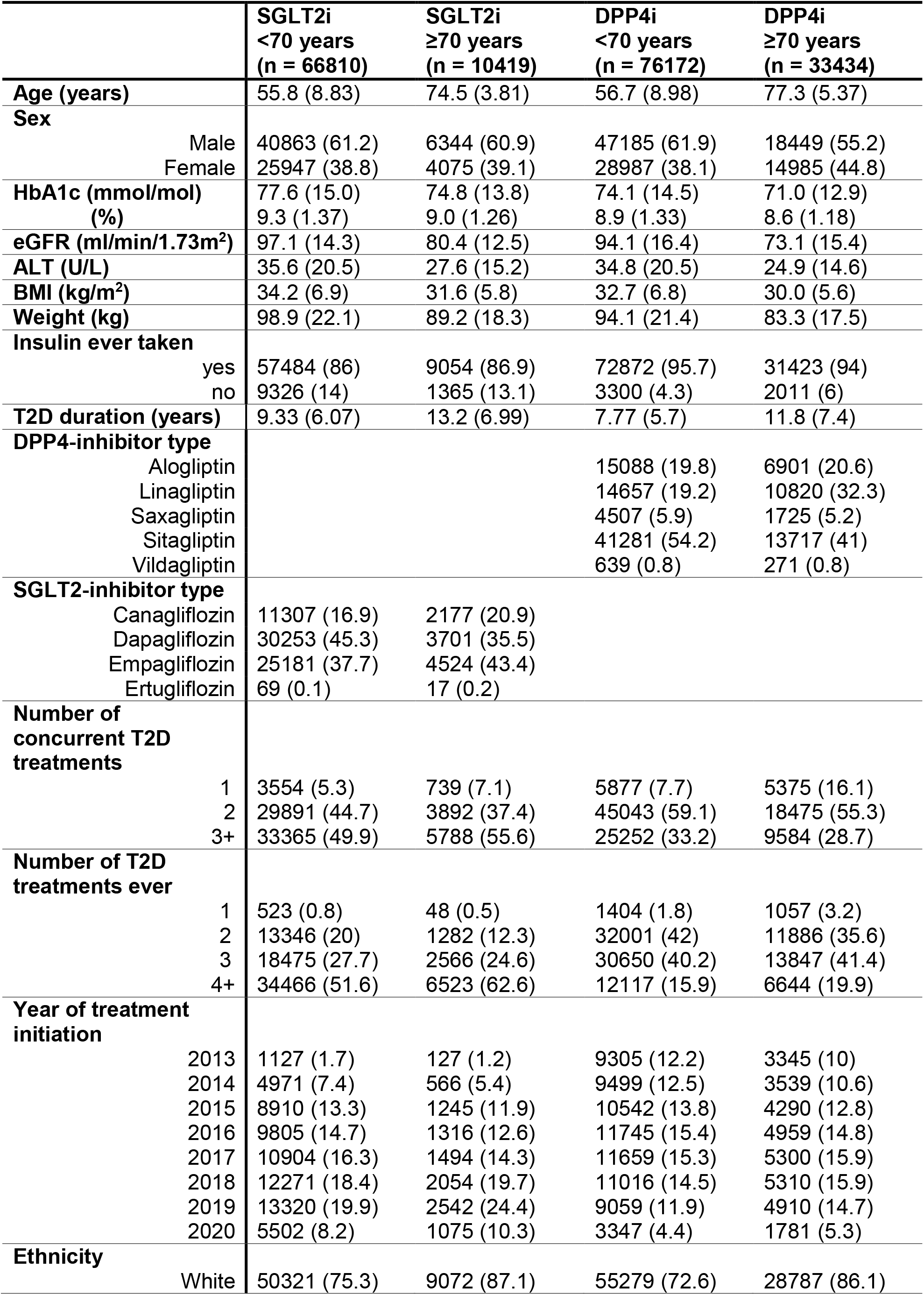
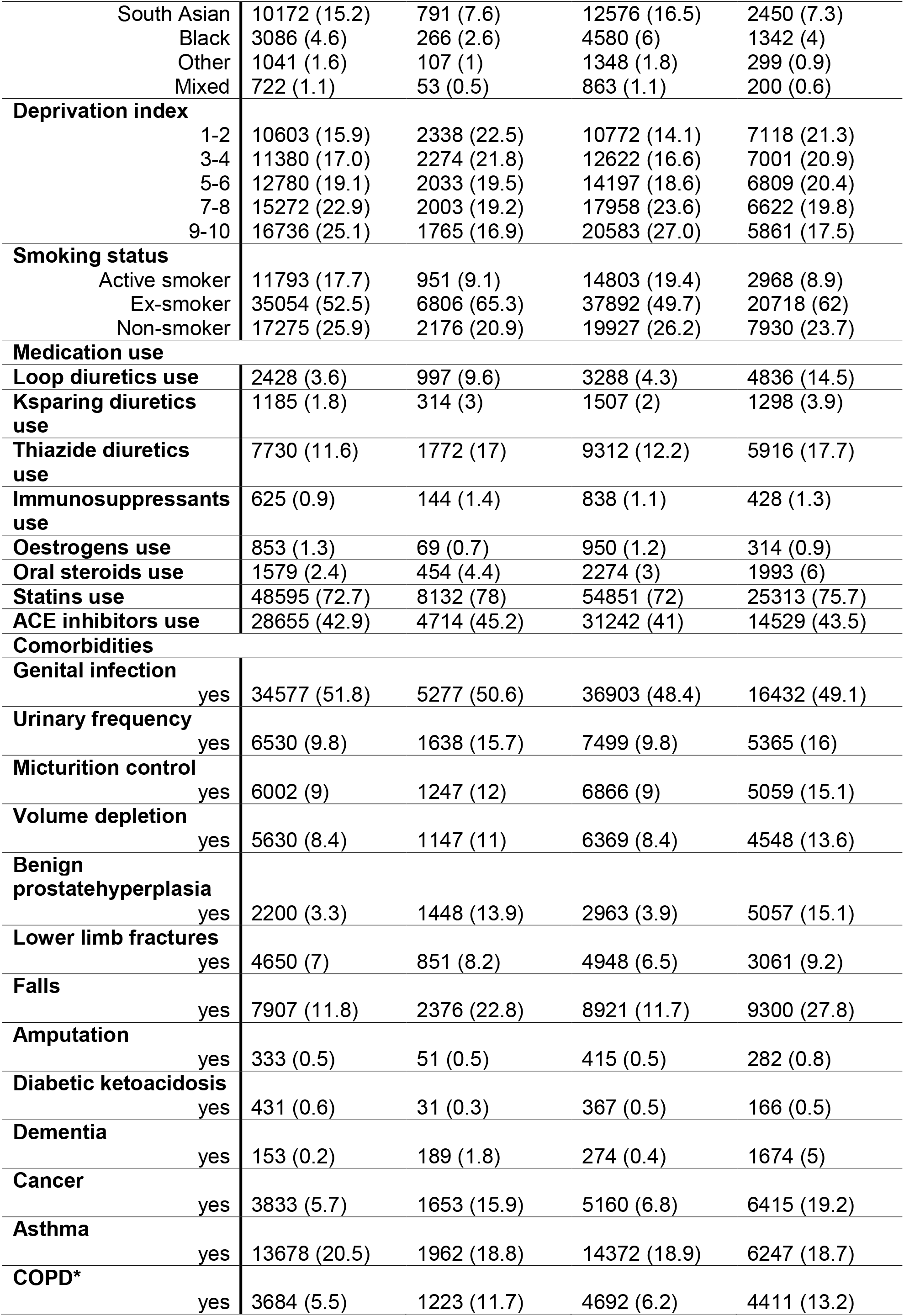
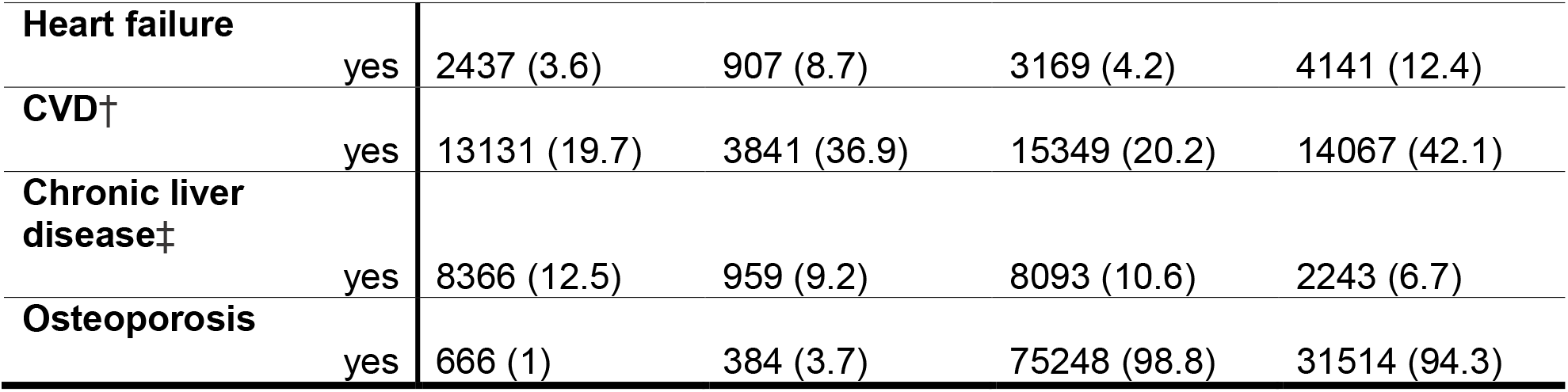
Baseline characteristics of the study cohort. Values for continuous variables are given in mean (standard deviation) and for binary and categorical variables in n (%). *COPD: chronic obstructive pulmonary disease, †CVD: composite of myocardial infarction, stoke, revascularisation, ischemic heart disease, angina, peripheral arterial disease, transient ischemic attack

### Statistical Methods

#### Causal analysis

When analysing treatment effects from observational data, bias due to confounding by indication is a major challenge. The confounding pre-treatment variables affect the outcome and the treatment decision simultaneously. As a result, it is possible that they differ in distribution between individuals who received the study and comparator treatment. [26] Traditional methods such as propensity score matching can mitigate the risk of bias by adjusting for measured confounders, but they cannot control for variables that are not recorded in the data, which can lead to unmeasured confounding. [26] With the Instrumental Variable (IV) approach and given a suitable instrument, treatment effects can be estimated in the presence of residual or unmeasured confounding without bias. [27] The basic idea of the IV approach is that a suitable IV is used to extract variation of the treatment that is free of unmeasured confounding. This variation is then utilized to estimate the treatment effect. [26] We employ the advanced IV approach proposed by Ertefaie et al. [28] which makes use of observed treatment behaviour and covariates to construct a proxy for prescription preference. Importantly, the method is capable of estimating the treatment effect without bias even in the presence of non-ignorable missingness in covariates. Our analysis did therefore not rely on a possibly selective complete case dataset. A more detailed explanation of this approach and a description of the assumed data structure for this study can be found in the supplementary material.

All binary AE outcomes were modelled using generalized Poisson regression with follow-up time (in days) as offset. For the estimation of the treatment effect of SGLT2-inhibitors on achieved HbA1c and weight a linear outcome model was used. Models used in the IV estimation and for all outcomes of interest were adjusted using different sets of relevant covariates specific to each outcome. A summary of all models is provided in the supplementary material in Supp. Table 1.

#### Sensitivity Analysis

We performed the following sensitivity analyses to assess the robustness of our findings: 1) To increase power due to the low number of events for several outcomes, we defined additional composite outcomes of osmotic symptoms (comprising volume depletion/dehydration, micturition control and urinary frequency) and combined falls and lower limb fracture (as not all falls might be coded in the CPRD data and lower limb fractures are often caused by falls. Our code list for lower limb fractures excludes fractures of the foot but includes hip) fractures of which 98% are caused by a fall [29]; 2) We additionally censored individuals who switched or added any other T2D treatments other than the study treatments over follow-up; 3) We repeated the analysis using 1 year maximum follow-up time for AE outcomes to assess short term risks; 4) We excluded the second drug exposure period for individuals who initiated both treatments over the study period.

## Results

The study cohort included 186835 episodes of participants commencing SGLT2i-inhibitors or DPP4-inhibitors from 161825 individuals (25010 initiated both treatments) (Figure 1). There were 142982 episodes included in the analysis for adults under 70 (<70) (n=76172 SGLT2-inhibitors, n=66810 DPP4-inhibitors) and 43853 episodes for adults 70 and older (≥70) (n=10419 SGLT2-inhibitors, n=33434 DPP4-inhibitors). Table 1 shows the baseline characteristics of the study population by treatment arm and age group. In the supplementary material a more detailed summary of comorbidity history is provided (Supp. Table 2) as well as a summary of the amount of missing data for each clinical characteristic (Supp. Table 3).

Incidence rate ratio estimate (IRR) for each AEs of interest are reported in Figure 2, and person-years and average follow-up time for all AEs are reported in the supplementary material in Supp. Table 4.

**Figure 2:**
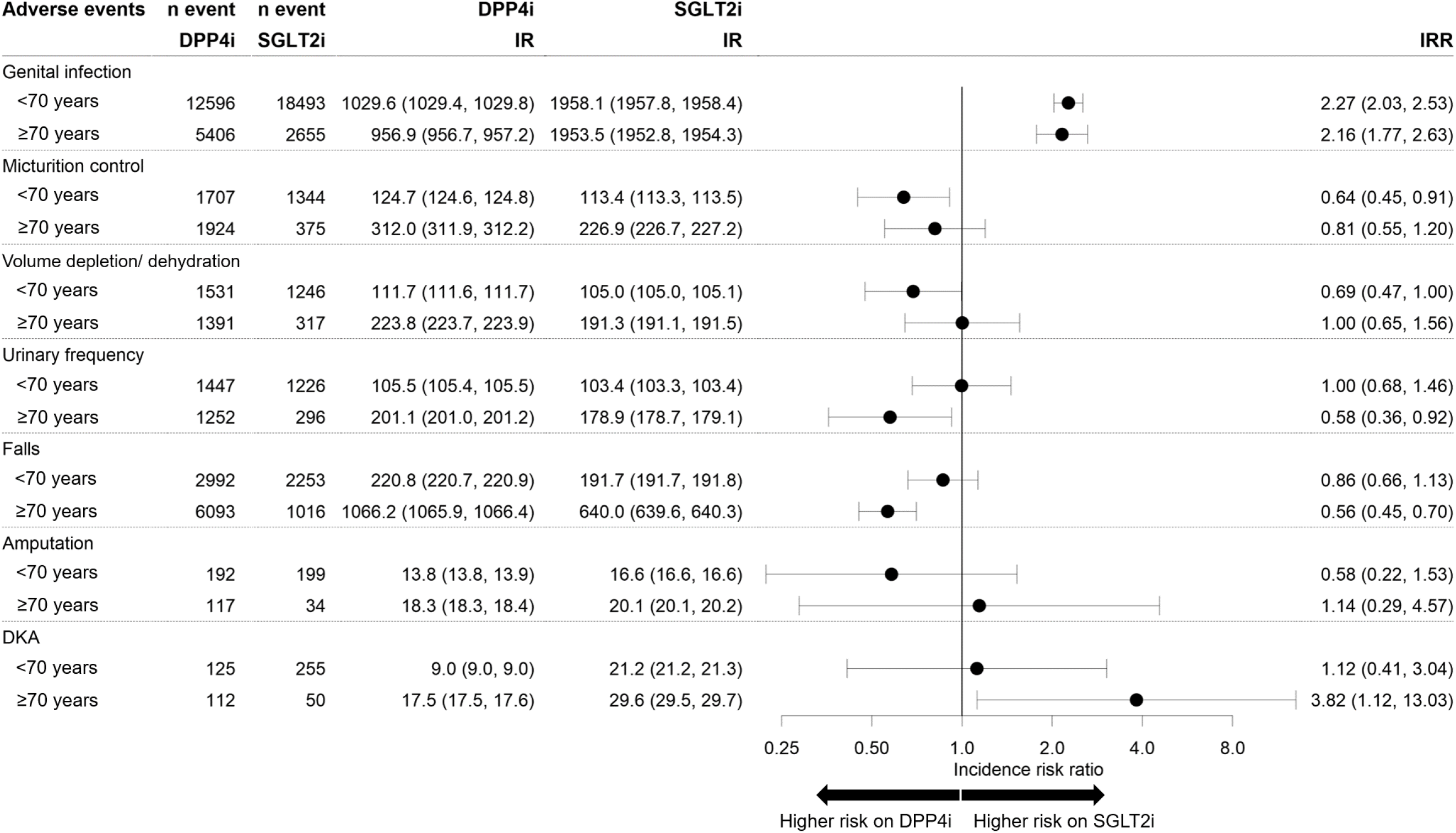
Causal effect estimation results of the incidence risk ratio (IRR) for adverse events estimated for n=142982 patients <70 (n=66810 SGLT2-inhibitors, n=76172 DPP4-inhibitors) and n=43853 ≥70 (n=10419 SGLT2-inhibitors, n=33434 DPP4-inhibitors). Additionally, the figure shows number (n) of events recorded and incidence rates (IR) per 10000 person-years. Values in brackets represent 95% confidence intervals.

### Risk of genital infection for people with T2D initiating SGLT2-inhibitors is similarly increased in adults under and over 70

Genital infections were the most commonly recorded AE (Figure 2), with the highest incidence in adults ≥70 initiating SGLT2-inhibitors (SGLT2-inhibitors incidence rate (IR) 1953.5 [95%CI 1952.8,1954.3] per 10000 person-years; DPP4-inhibitors 956.9 [95%CI 956.7, 957.2]). Causal treatment estimates suggested SGLT2-inhibitors were associated with a 2.16 (95%CI 1.77, 2.63) incidence rate ratio of genital infection compared with DPP4-inhibitors in adults ≥70, with a similar IRR in adults under 70 (2.27 [95%CI 2.03, 2.53].

### DKA is a rare AE and the risk increase with SGLT2-inhibitors may be restricted to adults over 70

DKA was rare event, and the highest incidence rate was recorded for ≥70 on SGTL2i (SGLT2-inhibitors IR 29.6 [CI 95% 29.5, 29.7] per 10000 person-years; DPP4-inhibitors 17.5 [CI 95% 17.5, 17.6]). Causal estimates suggested incidence risk ratio for DKA with SGLT2-inhibitors (compared to DPP4i) was increased for those ≥70 (IRR 3.82 [CI 95% 1.12, 13.03]), but not those under 70 (IRR 1.12 [CI 95% 0.41, 3.04]) (Figure 2).

### Risk of osmotic adverse events is not increased with SGLT2-inhibitors in adults under and over 70

Incidence rates for the AE micturition control for those ≥70 on SGLT2-inhibitors was 226.6 [CI 95% 226.7, 227.2] per 10000 person-years and the causal estimates did not show an increased risk in this patient group compared to people on DPP4-inhibitors (IRR: 0.81 [CI 95% 0.55, 1.20]). For the AE volume depletion (including dehydration) incidence rates in those ≥70 on SGLT2-inhibitors were 191.3 [CI 95% 191.1, 191.5] per 10000 person-years. Causal estimates of risk are not increased for this group (IRR: 1.00 [CI 95% 0.65, 1.56]). Additionally, the incidence rate of the AE urinary frequency was 178.9 [CI 95% 178.7, 179.1] per 10000 person-years and no increased risk was found for those ≥70 on SGLT2-inhibitors compared to those on DPP4-inhibitors (IRR: 0.58 [CI 95% 0.36, 0.92]) from the causal analysis.

### Risk of falls and amputations is not increased with SGLT2-inhibitors in adults under and over 70

The highest incidence rate for falls was recorded for those ≥70 (SGLT2-inhibitors IR 640.0 [CI 95% 639.6, 640.3] per 10000 person-years; DPP4-inhibitors 1066.2 [CI 95% 1065.9, 1066.4]). Results of the causal analysis did not show evidence of an increased incidence risk ratio of falls for SGLT2-inhibitors in comparison to DPP4-inhibitors treatment: IRR 0.86 [CI 95% 0.66, 1.13] for those <70 and 0.56 [CI 95% 0.45, 0.70] for those ≥70 (Figure 2).

Lower limb amputation was rare and a higher incidence rate was recorded for those ≥70 (SGLT2-inhibitors incident rate 18.3 [CI 95% 18.3, 1.84] per 10000 person-years; DPP4-inhibitors 20.1 [CI 95% 20.1, 20.2]). In causal analysis, there was no evidence of an increased risk of lower limb amputations (IRR 0.58 [CI 95% 0.22, 1.53] for those <70; 1.14 [CI 95% 0.29, 4.57] for those ≥70 (Figure 2).

### Glucose lowering efficacy of SGLT2-inhibitors is similar to DPP4-inhibitors in older patients, but in younger adults SGLT2-inhibitors are more effective

Unadjusted average HbA1c response for those <70 was −12.3 mmol/mol [CI 95% - 12.4, −12.1] (−1.1% [CI 95% −1.1, −1.1]) on SGLT2-inhibitors and −7.7 mmol/mol [CI 95% −7.8, −7.5] (−0.7% [CI 95% −0.7, −0.7]) on DPP4-inhibitors. For those ≥70, unadjusted HbA1c response was −9.9 mmol/mol [CI 95% −10.2, −9.5] (−0.9% [CI 95% −0.9, −0.9]) on SGLT2-inhibitors and −8.5 mmol/mol [CI 95% −8.7, −8.4] (−0.8% [CI 95% −0.8, −0.8]) on DPP4-inhibitors. Causal estimates for differences in HbA1c response and weight change between therapies are shown in Figure 3. For those <70, there was a greater reduction on in HbA1c with SGLT2-inhibitors compared to DPP4-inhibitors of −4 mmol/mol [CI 95% −4.8, −3.1] (−0.4% [CI 95% −0.4, −0.3]). For those ≥70, HbA1c response on both drug classes was similar (HbA1c differences between therapies −0.25 mmol/mol [CI 95% −1.63, 1.13], −0.02% [CI 95% −0.1, 0.1, favouring SGLT2-inhibitors). In contrast, the causal analysis results show a greater reduction in weight with SGLT2-inhibitors compared to DPP4-inhibitors in both age groups, with an SGLT2-inhibitors benefit of −2.6 kg [CI 95% −3, −2.3] for those <70 and −2.8 kg [CI 95% −3.3, −2.3] for those ≥70. Unadjusted average weight response was higher for patients initiating SGLT2i with −3.9 kg [CI 95% −4.0, −3.8] for those <70 and initiating SGLT2-inhibitors and −1.1 kg [CI 95% −1.1, −1.1] on DPP4-inhibitors respectively. For those ≥70, unadjusted weight response was −4.1 kg [CI 95% −4.2, −4.0] on SGLT2-inhibitors and −1.3 kg [CI 95% −1.3, −1.2] on DPP4-inhibitors.

**Figure 3:**
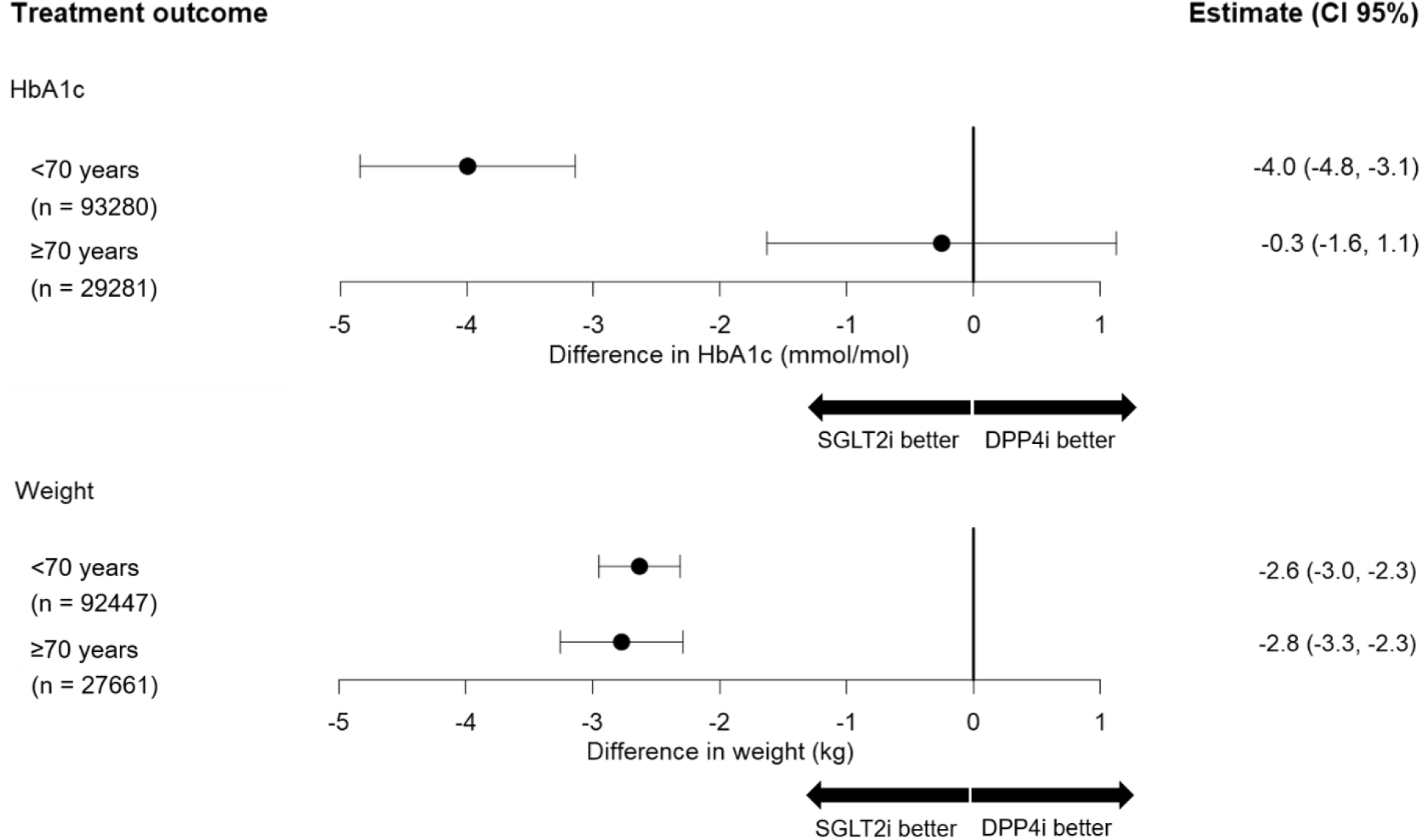
Causal effect estimation results for change in HbA1c (mmol/mol) and weight (kg).Point estimates represent the difference in outcome with SGLT2-inhibitors compared to DPP4-inhibitors, with negative values representing a greater HbA1c/weight reduction with SGLT2-inhibitors over DPP4-inhibitors. Numbers of n represent the cases with valid outcome value for which the complete case analysis is applied.

### Results of the sensitivity analysis are consistent with the main causal analysis results

Results of all sensitivity analyses are given in supplementary Supp. Table 5. Results were similar to the primary analysis when 1) using composite outcomes for osmotic symptoms and falls/ lower limb fractures; 2) censoring follow-up time at any change in treatment regime; 3) restricting maximum follow-up time post-drug initiation to one year (except that DKA risk in those ≥70 was no longer significantly increased); 4) excluding individuals initiating both treatments over the study period.

## Discussion

Our large-scale causal analysis provides important real-world evidence supporting careful use of SGLT2-inhibitors in older adults. Importantly, we found no increased risk of falls, osmotic symptoms, or amputations in those over 70. Adverse events of potential concern were genital infections and, rarely, DKA. We also demonstrate that SGLT2-inhibitors are effective in reducing HbA1c in this age group, although the substantially greater glucose lowering than DPP4-inhibitors in younger adults with this agent is absent in the elderly, where both agents had similar efficacy.

Risk of genital infections was increased on SGLT2-inhibitors to a similar degree in both those under and over 70. This finding complements similar findings in previous meta-analysis [30] and observational data [24], which did not specifically evaluate risk in older adults. Although we found DKA risk with SGLT2-inhibitors was elevated in those over 70, incidence was very low. This finding supports the warnings of the FDA [15] and the EMA [16] and stresses the need to take DKA risk factors into account when prescribing SGLT2-inhibitors to older people. [11, 17]

A greater average glycaemic efficacy with SGLT2-inhibitors compared to DPP4-inhibitors has been consistently shown in previous RCTs [31, 32], meta-analyses [33], and observational data [34] which did not specifically evaluate older adults. We identify heterogeneity in relative glycaemic efficacy, with greater efficacy in those <70 but not in those ≥70. This lack of glycaemic benefit with SGLT2-inhibitors in older adults may relate to the association between increasing age and lower eGFR, a known predictor of attenuated glycaemic response with SGLT2-inhibitors. [35] Weight reduction after SGLT2-inhibitor initiation is confirmed from our results for both age-stratified populations. Previous RCT meta-analysis results comparing SGLT2-inhibitors and DPP4-inhibitors showed a greater weight reduction with SGLT2-inhibitors of −2.45 kg [95% CI: −2.71, −2.19] [5]. The extent of weight reduction in our study is similar to these results.

A major strength of our causal analysis lies in the application of the advanced IV method by Ertefaie et al. [28], which addresses possible unmeasured confounding and does not rely on complete case analysis due to missingness in measured baseline characteristics. The analysis was conducted with a large real-world primary care dataset linked to hospitalization data, capturing a broad range of AEs for SGLT2-inhibitors with comprehensive primary and secondary care data.

Limitations of this study are that the analysis relies on correct clinical coding of the AEs, which can be subject to inaccuracies due to miscoding or non-coding. For example, some under-representation of genital infections might be possible as antifungal medication is available “over-the-counter” and can be treated without having presented to primary care. Additionally, information about the severity of the AEs was not available. [24] A limitation of the IV method is that some of the data structure assumptions made are not testable with the data. Additionally, as prescription preference was not measured in the data, our analysis relies on a proxy measurement, which might be subject to measurement errors. Previous similar IV analyses assessing relative effectiveness and risk of T2D treatments in the CPRD data have found that the IV assumptions are reasonable in this setting. [36, 37]

## Conclusion

SGLT2-inhibitors in older adults are effective and do not increase risk of dehydration, falls or urinary problems in older adults with T2D. However, risk of genital infections is increased, and DKA is a rare but severe adverse event of concern, meaning baseline DKA risk should be carefully assessed before initiation of SGLT2-inhibitors. This study provides a valuable causal analysis framework for the study of older adults who are generally not included in randomized controlled trials.

## Data Availability

CPRD data are available by application to the CPRD Independent Scientific Advisory Committee. R code to preproduce the analysis in this paper is available at https://github.com/Exeter-Diabetes/CPRD-Laura-SGLT2i-in-older-adults.

## Acknowledgements

This article is based on data from Clinical Practice Research Datalink obtained under licence from the UK Medicines and Healthcare products Regulatory Agency. CPRD data is provided by individuals and collected by the NHS as part of their care and support. Approval for the study was granted by the CPRD Independent Scientific Advisory Committee (ISCA 22_002000). This study was supported by the National Institute for Health and Care Research Exeter Biomedical Research Centre. The views expressed are those of the author(s) and not necessarily those of the NIHR or the Department of Health and Social Care.

## Funding and Assistance

This research was funded by the Medical Research Council (UK) (MR/N00633X/1), and supported by EFSD/Novo Nordisk. LMG, JMD, KGY and RH are supported by Research England’s Expanding Excellence in England (E3) fund. JMD is supported by the Wellcome Trust. ATH and BMS are supported by the NIHR Exeter Clinical Research Facility; the views expressed are those of the authors and not necessarily those of the NHS, the NIHR or the Department of Health. AGJ was supported by an NIHR Clinician Scientist fellowship (CS-2015-15-018).

## Conflict of Interest

APM received research funding from Eli Lilly and Company, Pfizer, and AstraZeneca. BMS are supported by the NIHR Exeter Clinical Research Facility. JB is a part time employee of Novo Nordisk. This project is unrelated to his work for the company. AGJ declares research funding to his university from the UK Medical Research Council, Diabetes, JDRF, and the European Foundation for the Study of Diabetes. Representatives from GSK, Takeda, Janssen, Quintiles, AstraZeneca and Sanofi attend meetings as part of the industry group involved with the MASTERMIND consortium. No industry representatives were involved in the writing of the manuscript or analysis of data. For all authors these are outside the submitted work; there are no other relationships or activities that could appear to have influenced the submitted work.

## Author Contributions and Guarantor Statement

LMG, APM, JMD, BMS and JB designed the study. APM, NT, AJ and AH provided valuable clinical insight and helped interpreting the results. KGY, RH, RC and APM developed code lists for the identification of relevant outcomes and comorbidities for the construction of the T2D cohort. KGY, RH, JMD and BMS constructed the T2D cohort of the CPRD data. From this cohort LMG identified individuals with relevant treatment regimes and characteristics for this study. LMG, JMD, BMS and JB developed the analysis strategy. LMG analysed the data under supervision of JMD, BS and JB. LMG drafted the original version of the paper which all authors helped to edit. All authors read and approved the final version of the manuscript. APM and JMD are the guarantors of this work and, as such, had full access to all the data in the study and takes responsibility for the integrity of the data and the accuracy of the data analysis.

## Prior Presentation

none

## Notes

### Competing Interest Statement

The authors have declared no competing interest.

