## Supplemental material for "Safety and effectiveness of SGLT2-inhibitors in people with type 2 diabetes over 70: UK population-based study using an Instrumental Variable approach"

### Online supplementary material

#### The Instrumental Variable approach as applied in this study

Supp. Figure 1 represents the assumed data structure of this observational study that are pertinent to the Instrumental Variable (IV) analysis. Arrows in the graph represent assumed causal relationships between the variables. The aim of the study is to estimate the causal effect of receiving SGLT2-inhibitors versus DPP4-inhibitors on the outcome(s) of interest. In particular, we assume that provider prescription preference is a suitable IV and fulfils the IV assumptions, conditional on a set of measured confounders, W. The IV assumptions are: (1) The IV must be strongly associated with the exposure given W, (2) be independent of unmeasured confounders given W and (3) not have a direct effect on the outcome of interest given W [1]. Necessary conditions for the IV assumptions to hold are that (1) between-provider variation in the use of study treatment exists, (2) individuals selection/assignment to a provider is unrelated to providers' preference of the study treatment, (3) a providers' use of treatment is independent of the use of alternative treatments that affect the outcome of interest. [2, 3].

As provider prescription preference is not directly measured in CPRD, and the set of measured confounders contains missing data, we use a proxy variable for it following the approach of Ertefaie et al. [4].

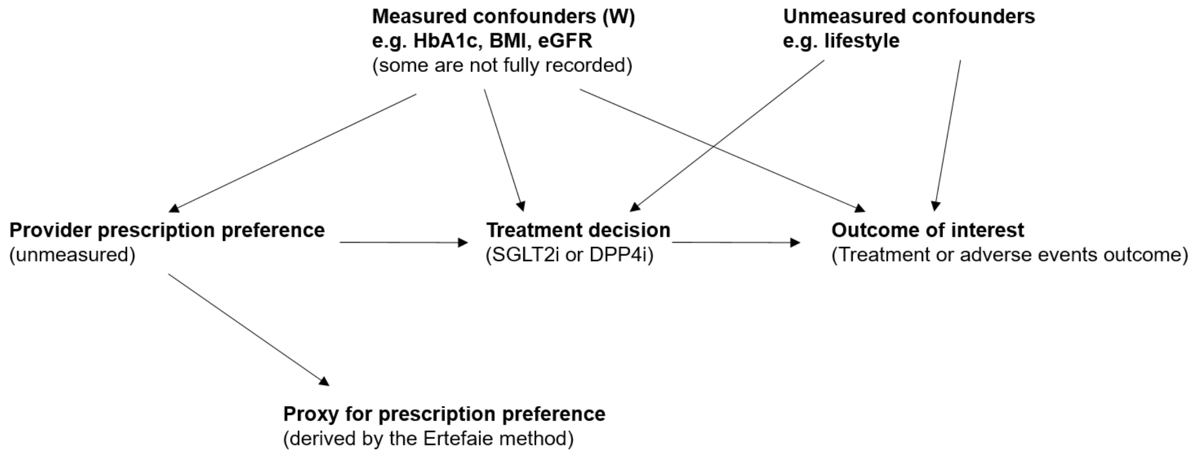


Supp. Figure 1: *Representation of assumed data structure in this study. Arrows in this plots indicate assumed relationships between variables. The missing of arrows indicates assumed lack of relationship. We assume that the variable provider prescription preference is a valid IV but it is not measured in the data at hand. Therefore, a proxy variable is derived using the IV approach by Ertefaie et al. 2017.*

The following non-technical description aims to give interested readers a better understanding of the steps necessary for the construction of a proxy variable for provider prescription preference and the estimation of the causal treatment effects. For a more in depth and mathematical description of the methods, we refer to the original paper by Ertefaie et al. [4].

The Ertefaie IV approach is conducted in two steps. To apply this approach the measured confounders/ covariates are grouped into:

- $W_{obs}$: all measured confounders that are observed for all individuals in the data and
- $W_{miss}$: all measured confounders with at least one missing data point.

Step 1 of the method aims to construct a binary proxy IV for provider prescription preference which will be used as instrument, taking the value 1 when a provider has a preference for SGLT2-inhibitors over DPP4-inhibitors, and 0 otherwise. It is constructed using a generalized mixed effect model for the treatment decision adjusted for all measured confounders ($W_{obs}$ and $W_{miss}$). The model is estimated using a complete case dataset (i.e. for individuals with complete information on $W_{obs}$ and $W_{miss}$) and with a random intercept for provider. From this model the fitted values of the random intercept and its empirical distribution is used for the construction of the instrument. The instrument will take on the value 1 for each provider with an estimated random intercept larger than the median of all estimated random intercepts, and 0 otherwise. Please note that the instrument can only be calculated for provider with at least one measured covariate completely measured ($W_{obs}$). If this is not the case, the provider will needs to be excluded from the IV analysis.

The second step of the Ertefaie method includes the calculation of the causal treatment effect with the Two-Stage Least Squares approach for continuous outcomes and the Two Stage Predictor Substitution method otherwise. [1] This estimation step is applied to all individuals in the dataset, but with adjustment for $W_{obs}$ only. Specifically, in stage 1, a logit model for the observed treatment decision is fitted which adjusts for $W_{obs}$ and the instrument. Thereafter, in stage 2, the outcome is regressed on the predicted treatment decision and $W_{obs}$ to estimate the causal treatment effect. For the continuous treatment outcomes, achieved HbA1c and weight, a linear outcome model is estimated. In case of binary adverse event outcomes, we used a Poisson model with follow-up time (in days) as offset.

| | Achieved HbA1c | Achieved weight | Genital infections | Osmotic symptoms | Falls | Lower limb amputations | Amputations | Diabetic ketoacidosis |
|---|---|---|---|---|---|---|---|---|
| **General characteristics and treatment regime information** | Age, sex, ethnicity, deprivation index, smoking status, diabetes duration, year of study treatment initiation, line of therapy, number of concurrent treatments, insulin taken | | | | | | | |
| **Biomarkers** | HbA1c, eGFR, BMI/ weight, ALT | | | | | | | |
| **History of comorbidities** | | | Genital infections | Osmotic symptom, benign prostate hyperplasia | Oestrogens, oral steroids, statins, Ksparing, loops, thiazide diuretics, ACE inhibitors | | | |
| **Additional treatments** | | | Immuno-suppressants, oestrogens, oral steroids | Ksparing, Loops, thiazide diuretics | lower limb fracture, falls, amputation, Diabetic ketoacidosis, Dementia, Cancer, Asthma, COPD*, heart failure, CVD †, CLD ‡, osteoporosis | | | |

Supp. Table 1: Summary of the relevant covariates for each outcome of interest used in the IV estimation. For the analysis of achieved weight, baseline weight instead of baseline BMI was included. Models of the first step of IV method by Ertefaie et al. 2017 and the first stage regression model of the IV estimation adjusted for all relevant covariates, whereas the outcome models only included completely recorded covariates. *COPD: chronic obstructive pulmonary disease, †CVD: composite of myocardial infarction, stoke, revascularisation, ischemic heart disease, angina, peripheral arterial disease, transient ischemic attack, ‡CLD: chronic liver disease.

| | SGLT2i <70 years (n = 66810) | SGLT2i ≥70 years (n = 10419) | DPP4i <70 years (n = 76172) | DPP4i ≥70 years (n = 33434) |
|---|---|---|---|---|
| **Genital infection** | | | | |
| Never | 32233 (48.2) | 5142 (49.4) | 39269 (51.6) | 17002 (50.9) |
| <1 year | 8950 (13.4) | 1197 (11.5) | 10566 (13.9) | 3920 (11.7) |
| 1-5 years | 13114 (19.6) | 1890 (18.1) | 13717 (18) | 5610 (16.8) |
| >5 years | 12513 (18.7) | 2190 (21) | 12620 (16.6) | 6902 (20.6) |
| **Urinary frequency** | | | | |
| Never | 60280 (90.2) | 8781 (84.3) | 68673 (90.2) | 28069 (84) |
| <1 year | 819 (1.2) | 178 (1.7) | 1170 (1.5) | 832 (2.5) |
| 1-5 years | 2309 (3.5) | 598 (5.7) | 2802 (3.7) | 1969 (5.9) |
| >5 years | 3402 (5.1) | 862 (8.3) | 3527 (4.6) | 2564 (7.7) |
| **Micturition control** | | | | |
| Never | 60808 (91) | 9172 (88) | 69306 (91) | 28375 (84.9) |
| <1 year | 786 (1.2) | 180 (1.7) | 1087 (1.4) | 1024 (3.1) |
| 1-5 years | 2024 (3) | 425 (4.1) | 2340 (3.1) | 1732 (5.2) |
| >5 years | 3192 (4.8) | 642 (6.2) | 3439 (4.5) | 2303 (6.9) |
| **Volume depletion** | | | | |
| Never | 61180 (91.6) | 9272 (89) | 69803 (91.6) | 28886 (86.4) |
| <1 year | 730 (1.1) | 164 (1.6) | 960 (1.3) | 815 (2.4) |
| 1-5 years | 1881 (2.8) | 398 (3.8) | 2173 (2.9) | 1551 (4.6) |
| >5 years | 3019 (4.5) | 585 (5.6) | 3236 (4.2) | 2182 (6.5) |
| **Falls** | | | | |
| Never | 58903 (88.2) | 8043 (77.2) | 67251 (88.3) | 24134 (72.2) |
| <1 year | 1128 (1.7) | 603 (5.8) | 1473 (1.9) | 2989 (8.9) |
| 1-5 years | 2738 (4.1) | 902 (8.7) | 3224 (4.2) | 3634 (10.9) |
| >5 years | 4041 (6) | 871 (8.4) | 4224 (5.5) | 2677 (8.0) |
| **Amputation** | | | | |
| Never | 66477 (99.5) | 10368 (99.5) | 75757 (99.5) | 33152 (99.2) |
| <1 year | 75 (0.1) | 6 (0.1) | 111 (0.1) | 62 (0.2) |
| 1-5 years | 147 (0.2) | 20 (0.2) | 182 (0.2) | 115 (0.3) |
| >5 years | 111 (0.2) | 25 (0.2) | 122 (0.2) | 105 (0.3) |
| **Diabetic ketoacidosis** | | | | |
| Never | 66379 (99.4) | 10388 (99.7) | 75805 (99.5) | 33268 (99.5) |
| <1 year | 61 (0.1) | 6 (0.1) | 95 (0.1) | 66 (0.2) |
| 1-5 years | 173 (0.3) | 15 (0.1) | 129 (0.2) | 48 (0.1%) |
| >5 years | 197 (0.3) | 10 (0.1) | 143 (0.2) | 52 (0.2) |

Supp. Table 2: Detailed description of potential recurrent comorbidities in the study population recorded prior to study treatment initiation.

| | SGLT2i<br><70 years<br>(n = 66810) | SGLT2i<br>≥70 years<br>(n = 10419) | DPP4i<br><70 years<br>(n = 76172) | DPP4i<br>≥70 years<br>(n = 33434) |
|---|---|---|---|---|
| **HbA1c (mmol/mol and %)** | 8973 (13.4) | 1406 (13.5) | 7559 (9.9) | 3999 (12) |
| **eGFR (ml/min/1.73m$^2$)** | 322 (0.5) | 25 (0.2) | 555 (0.7) | 170 (0.5) |
| **ALT (U/L)** | 4206 (6.3) | 577 (5.5) | 5354 (7.0) | 2158 (6.5) |
| **BMI (kg/m$^2$)** | 2646 (4) | 371 (3.6) | 3701 (4.9) | 1988 (5.9) |
| **Weight (kg)** | 1496 (2.2) | 215 (2.1) | 2370 (3.1) | 1440 (4.3) |

Supp. Table 3: Summary of missing data in baseline characteristics of the study population values are given in absolute frequencies in n (%).

| | Person-years of follow-up | Average follow-up time (years) |
|---|---|---|
| **Adverse event** | | |
| Genital infections | 286867.4 | 1.54 (1.03) |
| Volume depletion | 334448 | 1.79 (1) |
| Micturition control | 333607.1 | 1.79 (1) |
| Urinary frequency | 334596.2 | 1.79 (1) |
| Falls | 326040.4 | 1.75 (1) |
| Amputation | 339284.1 | 1.82 (0.99) |
| DKA | 339574.1 | 1.82 (0.99) |
| **Composite adverse event** | | |
| Osmotic symptoms | 329345.2 | 1.76 (1) |
| Falls + lower limb fractures | 339848.6 | 1.82 (0.99) |

Supp. Table 4: Person-years of follow-up calculated as the total follow-up time and average follow-up time (in years) of the adverse events. Values for the average follow-up time are shown as mean (standard deviation).

| | Composite outcomes | Censoring scheme | Follow-up time | Second study period |
|---|---|---|---|---|
| **Treatment outcomes**<br>**Average relative difference (CI 95%)** | | | | |
| HbA1c (mmol/mol, %) | | | | |
| <70 years<br>≥70 years | | | | -4.3 (-5.1, -3.5), 0.4 (-0.5, -0.3)<br>-0.3 (-1.8, 1.2), 0.03 (-0.2, 0.1) |
| Weight (kg) | | | | |
| <70 years<br>≥70 years | | | | -2.7 (-2.9, -2.3)<br>-2.7 (-3.2, -2.2) |
| **Adverse event outcomes**<br>**IRR (CI 95%)** | | | | |
| Genital infection | | | | |
| <70 years<br>≥70 years | | 2.42 (2.14, 2.74)<br>2.16 (1.74, 2.68) | 2.55 (2.24, 2.9)<br>2.0 (1.59, 2.56) | 2.11 (1.9, 2.34)<br>2.04 (1.66, 2-51) |
| Micturition control | | | | |
| <70 years<br>≥70 years | | 0.69 (0.46, 1.02)<br>0.91 (0.58, 1.42) | 0.72 (0.45, 1.16)<br>0.91 (0.54, 1.51) | 0.73 (0.52, 0.98)<br>1.03 (0.68, 1.54) |
| Volume depletion/ dehydration | | | | |
| <70 years<br>≥70 years | | 0.73 (0.48, 1.11)<br>1.1 (0.67, 1.82) | 0.8 (0.49, 1.32)<br>1.33 (0.74, 2.38) | 0.73 (0.52, 1.03)<br>1.35 (0.85, 2.14) |
| Urinary Frequency | | | | |
| <70 years<br>≥70 years | | 0.99 (0.64, 1.52)<br>0.57 (0.33, 0.97) | 0.95 (0.57, 1.58)<br>0.75 (0.4, 1.39) | 1.33 (0.94, 1.88)<br>0.57 (0.34, 0.94) |
| Osmotic symptoms (composite) | | | | |
| <70 years<br>≥70 years | 0.81 (0.62, 1.06)<br>0.82 (0.6, 1.12) | | | |
| Falls | | | | |
| <70 years<br>≥70 years | | 0.83 (0.61, 1.13)<br>0.62 (0.48, 0.8) | 0.89 (0.61, 1.3)<br>0.46 (0.34, 0.61) | 0.95 (0.74, 1.2)<br>0.62 (0.49, 0.79) |
| Falls/ lower limb fractions (composite) | | | | |
| <70 years<br>≥70 years | 0.88 (0.68, 1.13)<br>0.59 (0.48, 0.74) | | | |
| Amputation | | | | |
| <70 years<br>≥70 years | | 0.91 (0.26, 3.17)<br>0.72 (0.1, 5.1) | 0.22 (0.05, 0.92)<br>1.4 (0.17, 11.79) | 0.84 (0.36, 2.0)<br>1.08 (0.26, 4.53) |
| DKA | | | | |
| <70 years<br>≥70 years | | 2.5 (0.37, 17.0)<br>12.05 (1.6, 91.34) | 2.46 (0.54, 11.19)<br>1.7 (0.3, 9.63) | 1.8 (0.72, 4.53)<br>6.19 (1.78, 21.5) |

Supp. Table 5: Summary of the sensitivity analysis results. Results for treatment outcomes are given in average relative difference in outcome measure and AE results as presented as incidence risk ratio. All results are shown with 95% confidence intervals. Composite outcomes: Analysis using a composite for osmotic symptoms as well as falls and lower limb fractures. Censoring scheme: censoring for the AE is done in case of any change of the treatment regime. Follow-up time: Follow-up time of the AEs was maximum 1 year. Second study period: Exclusion of the second study period for individuals who initiated both treatments.
